# Breast cancer is linked to changes in the urinary extracellular vesicle proteome

**DOI:** 10.64898/2026.05.08.26352674

**Authors:** Noura Laziri, Nur Aimi Aliah Zainurin, Anuradha U K H Bambarandhage, Olugbenga Samuel Babatunde, Tim Gate, Helen Tench, Daxu Fu, Xin Zhang, Manfred Beckman, Helen Phillips, Mandana Pennick, Russell M. Morphew, Luis A J Mur

## Abstract

Breast cancer (BC) remains a leading cause of morbidity and mortality worldwide. Early detection remains the most effective strategy for improving prognosis. We explored the urinary extracellular vesicle (uEV) proteome for changes linked to BC which could also be potential biomarkers. Urine samples were collected from 20 participants across four groups (n = 5 each): newly diagnosed BC patients, benign breast disease (BBD) patients, individuals with breast cancer symptoms (symptom control, SC), and age-matched healthy controls (HC). EVs were isolated using size exclusion chromatography and extracted proteins were analysed using a GeLC proteomic approach. Proteins were identified and quantified using Proteome Discoverer and further analysed using MetaboAnalystR, Funrich and Metascape. A total of 256 proteins were identified from the uEV preparations. BC comparisons with BBD, SC and HC identified 7 proteins differentially expressed proteins (DEP); SERPINB1 — Serpin family B member 1, LCN1 — Lipocalin 1, SIRPA — Signal regulatory protein alpha, ACTB — Actin, beta, YWHAZ —Tryptophan 5-monooxygenase activation protein zeta, Ig JCHAIN and APOA1 — Apolipoprotein A1. Receiver Operator Characteristic (ROC) curve assessments suggested that each DEP protein had an area under the curve (AUC) of > 0.8. These findings highlight EV-derived proteins as promising non-invasive biomarkers for breast cancer detection, warranting further validation in larger cohorts.

## Introduction

Breast cancer (BC) represents a major global health problem. In 2022, BC was the most diagnosed cancer in women in 157 countries (World Health Organisation, 2025). However, current diagnostic and screening methods are often invasive, costly, and show poorer sensitivity for early-stage detection (Bhushan et al., 2021). This highlights the urgent need for alternative, non-invasive approaches that can improve early diagnosis and patient outcomes.

BC development and progression are driven by a multitude of cellular and molecular alterations that reshape the tumour microenvironment. Well-established genetic mutations in TP53 and BRCA1/2 are important in cell cycle and DNA repair dysregulation in BC (Greenblatt et al., 2001; Zhao et al., 2013). Also important are changes in the extracellular matrix (ECM) which is a defining feature, influencing tumour growth, invasion, and metastasis (Chen et al., 2025; Li et al., 2024). Metalloproteinases (MMPs) are the key mediators of ECM remodelling, degrading the structural components of the matrix, thereby facilitating tissue remodelling, invasion, and cell migration. Epithelial Mesenchymal Transition (EMT), a process marked by the loss of E-cadherin and the gain of N-cadherin, enables cells to acquire a motile and invasive mesenchymal phenotype. These changes are often associated with the upregulation of genes encoding MMPs and EMT regulators like TWIST and SNAIL (Jena & Janjanam, 2018). Collectively, these molecular and genetic changes highlight the critical role of ECM remodelling in breast cancer progression, driving invasion, metastasis, and shaping the tumour microenvironment. These could provide potential biomarkers for the initiation and development of BC.

As a potential source of such biomarkers, urine has emerged as a promising diagnostic biofluid due to its abundance, ease of collection, and stability, making it ideal for non-invasive disease detection (Balhara et al., 2023). Urine is rich in proteins, nucleic acids, lipids, and metabolites; biomolecules that can serve as potential biomarkers (Sequeira-Antunes & Ferreira, 2023). Our previous work has demonstrated urinary proteins associated with BC (Zainurin et al., 2025) and here we explore if these proteins are associated with urinary extracellular vesicles (uEVs).

EVs are lipid bilayer vesicles secreted by most cells, and especially from cancerous cells, and are found in all body fluids including urine (Doyle & Wang, 2019). Within the context of cancer, EVs transfer oncogenic proteins, RNAs, and metabolites that promote tumour growth, invasion, metastasis and immune evasion. Thus, EVs reshape the tumour microenvironment and prepare pre-metastatic niches (Greening et al., 2025). Importantly, studies have revealed a dynamic bidirectional relationship between EVs and the ECM. ECM properties such as stiffness and hypoxia influence EV biogenesis, cargo composition (proteins and miRNAs), and release, thereby promoting tumour progression (Peppicelli et al., 2025; Wang et al., 2025). Conversely, EVs can directly remodel the ECM by delivering matrix metalloproteinases (MMPs) and other enzymes that degrade structural components, facilitating invasion and metastasis (Shimoda & Khokha, 2017).

Herein, we assess if EV-derived proteins differ in samples from women with BC compared to those with benign breast disease (BBD) or were symptomatic but non-diseased controls (SC) and healthy controls (HC). This study identified key protein changes in BC which could serve as potential biomarkers for BC and suggest key systemic changes in BC patients.

## Materials and Methods

### Ethics approval and recruitment criteria

Ethical approval for the BECA (“OMICs Approaches to Improve the Diagnosis, Management and Treatment of Breast Cancer “) clinical project was obtained from the Health Research Authority (HRA) and Health and Care Research Wales (HCRW) IRAS Project ID: 306872 and CPMS Study ID: 54143. Female adult participants (aged over 18 years) attending the rapid access clinic at West Hertfordshire Hospital (UK) and presenting with symptoms suggestive of BC were recruited for the study. Subsequently, clinical assessments indicated that recruits either had BC, BBD or were symptom controls (SC). The SC group consisted of women who presented with breast related symptoms, such as breast pain and breast lumps, but whose clinical examination and imaging confirmed the absence of both BC and BBD. BECA recruits were also obtained from volunteers from Aberystwyth University (UK) with no diagnosis or history of breast disease and these were considered as healthy controls (HC). Informed consent was obtained from all individuals prior to participation in the study. Five samples were randomly selected from these cohorts for assessments of uEV proteomes (Table 1).

**Table 1.** Patients’ Demographic characteristics.

| Sample groups |  | Breast cancer (BC) | Benign breast disease (BBD) | Symptom control (SC) | Healthy control (HC) | Total |
| --- | --- | --- | --- | --- | --- | --- |
| Number of participants |  | 5 | 5 | 5 | 5 | 20 |
| Age range (Median ± SD) | Overall | 28-60<br>(51± 12.05) | 34-70<br>(47± 14.58) | 32-70<br>(50 ± 3) | 44-52<br>(54± 14.71) | 28-70<br>(50± 10.86) |
| Number of participants by age range, n (%) | <40 | 1 (20) | 2 (40) | 0 (0) | 1 (20) | 4 (20) |
|  | 41-60 | 4 (80) | 2 (40) | 5 (100) | 3 (60) | 14 (70) |
|  | 61-80 | 0 (0) | 1 (20) | 0 (0) | 1 (20) | 2 (10) |
|  | >81 | 0 (0) | 0 (0) | 0 (0) | 0 (0) | 0 (0) |
|  | Not stated | 0 (0) | 0 (0) | 0 (0) | 0 (0) | 0 (0) |
| BMI (kg/m<sup>2</sup>) (Mean ± SD) | | 24.07(±4.4) | 25.78 (±3.43) | 24.5 (±4.92) | 22.69 (±5.25) | 24.07 (±4.15) |
| Smoking habits, n (%)<br>(n=36) | Current | 0 (0) | 0 (0) | 0 (0) | 0 (0) | 0 (0) |
|  | Ex | 1 (20) | 1 (20) | 2 (40) | 0 (0) | 4 (20) |
|  | Never | 4 (80) | 4 (80) | 3 (60) | 5 (100) | 16 (80) |
| History of Cancer, (%) |  | 1 (20) | 0 (0) | 3 (60) | (0) | 4 (20) |
| Family history with cancers (%) |  | 2 (40) | 3 (60) | 5 (100) | 2 (40) | 12 (60) |

### Sample preparation and uEV extraction

Urine samples were centrifuged at 365 × *g* (Multicentrifuge 3SR, Heraeus Kendro, Germany), and the EV-containing supernatant retained and stored at −80°C prior to uEV extraction. Upon thawing, protease inhibitors (cOmplete™ Mini, EDTA-free, catalogue 11836170001) were added to the urine samples (20 µl per each 1 ml). All urine samples were filtered using a 0.22 µm syringe filter and concentrated using an Amicon® Ultra Centrifugal Filter (10 kDa MWCO), by centrifugation (High Speed Refrigerated Micro Centrifuge, DLAB Scientific Co., Ltd., Zhenjiang, China) at 4000 × *g* for 20 min. The filtrate was stored at −20°C. uEV fractionation was performed using qEVoriginal columns and an Automatic Fraction Collector (AFC; Izon Science Ltd., Christchurch, New Zealand) according to the manufacturer’s instructions. Fractions were collected in 1.5 mL microcentrifuge (Greiner Bio-One, Germany), and the first three EV-rich fractions were pooled and concentrated using Amicon® Ultra Centrifugal Filter (10 kDa MWCO) to give a final uEV volume of 50 µl.

### Transmission electron microscopy

The isolation of uEVs was confirmed by transmission electron microscope (TEM). Aliquots of 10 µL of isolated uEV suspension was added to Formvar/Carbon coated grids (CN Technical, UK) and was left to absorb for 45 min. Grids were transferred to 10 µL of 4% w/v uranyl acetate for 5 min and visualised after 24 hours using JEOL JEM1010 transmission electron microscope (JEOL Ltd, Japan).

### GeLC Proteomics

Sodium Dodecyl Sulfate–Polyacrylamide Gel Electrophoresis (SDS-PAGE) was based on the protocol of (Brunelle & Green, 2014). Extracted uEV proteins were denatured in SDS sample buffer at 95 °C, briefly sonicated, and centrifuged at 10,000 rpm for 10 min prior to separation of the supernatants on a 12.5% polyacrylamide gel. For in-gel digestion, uEV protein samples were run until the loading dye (0.1 % bromophenol blue, 50 % glycerol) front reached 0.5 cm from the top of the gel. The 0.5 cm gel segments corresponding to each well were excised and placed in 0.5 mL microcentrifuge tube (Greiner Bio-One, Germany) prior to in gel digestion. Gel pieces were dehydrated with 100% acetonitrile (ACN) for 15 min at 37°C. The ACN was then discarded, and the gel pieces were dried at 50°C with the lids open. Then, 100 µl of 10 mM dithiothreitol (DTT) was added, and the gel pieces were left to incubate for 30 minutes at 80°C. The liquid was discarded and 100 µl of 55 mM iodoacetamide (IAA) was added and the tubes were left to incubate for 20 min at room temperature. The liquid was discarded once again, and the gel pieces were washed with 50 mM NH₄HCO₃ (pH 8) and 50% ACN for 15 min at room temperature. Next, 100% ACN was added to dehydrate the gel pieces for 15 minutes at room temperature and then dried with the microcentrifuge lids open at 50°C for 30 min. The gel pieces were then left to incubate overnight in 50 mM NH₄HCO₃ containing trypsin (10 ng/µl). The incubation time did not exceed 16 h.

Subsequently, 20–50 µl of ddH₂O was added to the gel pieces and the tubes were vortexed and centrifuged at 5000 × *g*. The resulting eluate was transferred to new tubes. To the gel pieces remaining in the original tubes, 50 µL of 50% ACN with 5% formic acid was added, and the tubes were shaken for 1 hour at room temperature, followed by brief centrifugation. This second eluate was collected and combined with the previously recovered liquid. The combined supernatant was dried in a speed vacuum (VACUUBRAND, Germany) and stored at –20°C. Immediately before mass spectrometry, all pellets were re-suspended in 20 µL of 0.1% formic acid.

### Mass Spectrometry Analysis

Protein samples were analyzed by LC-MS/MS using an Orbitrap Fusion Tribrid mass spectrometer (Thermo Scientific) coupled to an UltiMate 3000 RSLCnano system with an EASY-Spray source and C18 nano column (75 μm × 750 mm, 2 μm particle size). Peptides were separated under a gradient of 0.1% formic acid in water and acetonitrile at a flow rate of 200 nL/min. Data acquisition was performed in positive ion mode with a spray voltage of 1.8 kV and ion transfer temperature of 275°C. Full MS scans were recorded in the Orbitrap at 120,000 resolution across m/z 375–1500, followed by data-dependent MS/MS for precursors with charge states 2–7 using collision-induced dissociation at 35% energy. Fragment ions were detected in the ion trap, and dynamic exclusion was applied to minimize repeated sequencing.

### Proteome Discoverer

Proteomic data was assessed using Proteome Discoverer (Thermo Scientific, USA), version 2.5.0.400 using MASCOT and SEQUEST HT against the GPM-cRAP (common contaminants) and GRCH38 (human reference proteome) databases. The digesting enzyme was set to Trypsin, allowing a maximum of two missed cleavages, and peptide length range 5 - 144 amino acids. The precursor mass tolerance was set at 10 ppm, while the fragment mass tolerance was 0.02 Da. For quality control, a target false discovery rate (FDR) threshold of 0.01 was applied for filtering both peptides and proteins.

### Statistical Analysis

MetaboAnalystR 6.0 (https://www.metaboanalyst.ca/), an R-based platform for comprehensive statistical and functional analysis of high-dimensional omics data (Pang et al., 2024), was used to visualise and analyse the proteomic datasets based on principal component analyses (PCA), partial least squared – discriminant analyses (PLS-DA) and heat maps. Data were normalised using log₁₀ transformation and Pareto scaling prior to statistical evaluation. The FunRich tool (http://www.funrich.org/) was used to visualise overlapping proteins and explore their association with extracellular vesicles (Pathan et al., 2015). Enrichment analysis was performed using Metascape (https://metascape.org/) to identify significantly enriched pathways and biological processes (Zhou et al., 2019).

## Results

### Assessments of the whole urinary proteome

In our previous study we conducted an extensive (n= 130) study of proteomic changes in urine linked to BC (Zainurin et al., 2025). In this current study, we wish to assess how far these changes represented changes in the uEV proteome.

We assessed the whole urinary proteins within urine samples (BC, BBD, SC and HC; n = 5 in each class) which would subsequently be used to determine the uEV proteome. This would allow direct comparisons between the whole urine and uEV proteome. The derived results were assessed via a series of pairwise comparisons (BC vs BBD, BC v SC and BC vs. HC) were undertaken based on volcano plots (*P* <0.05/ fold change ≥ 2) (Table S1). The outputs were compared using a Venn Diagram (Figure 1a). This indicated 49 differentially expressed proteins (DEP) were common to all comparisons. It should be noted that there were 104 DEP (the largest number) specific to the BC vs. BBD comparison that would not have been observed if only SC and/or HC samples were considered.

**Figure 1:**
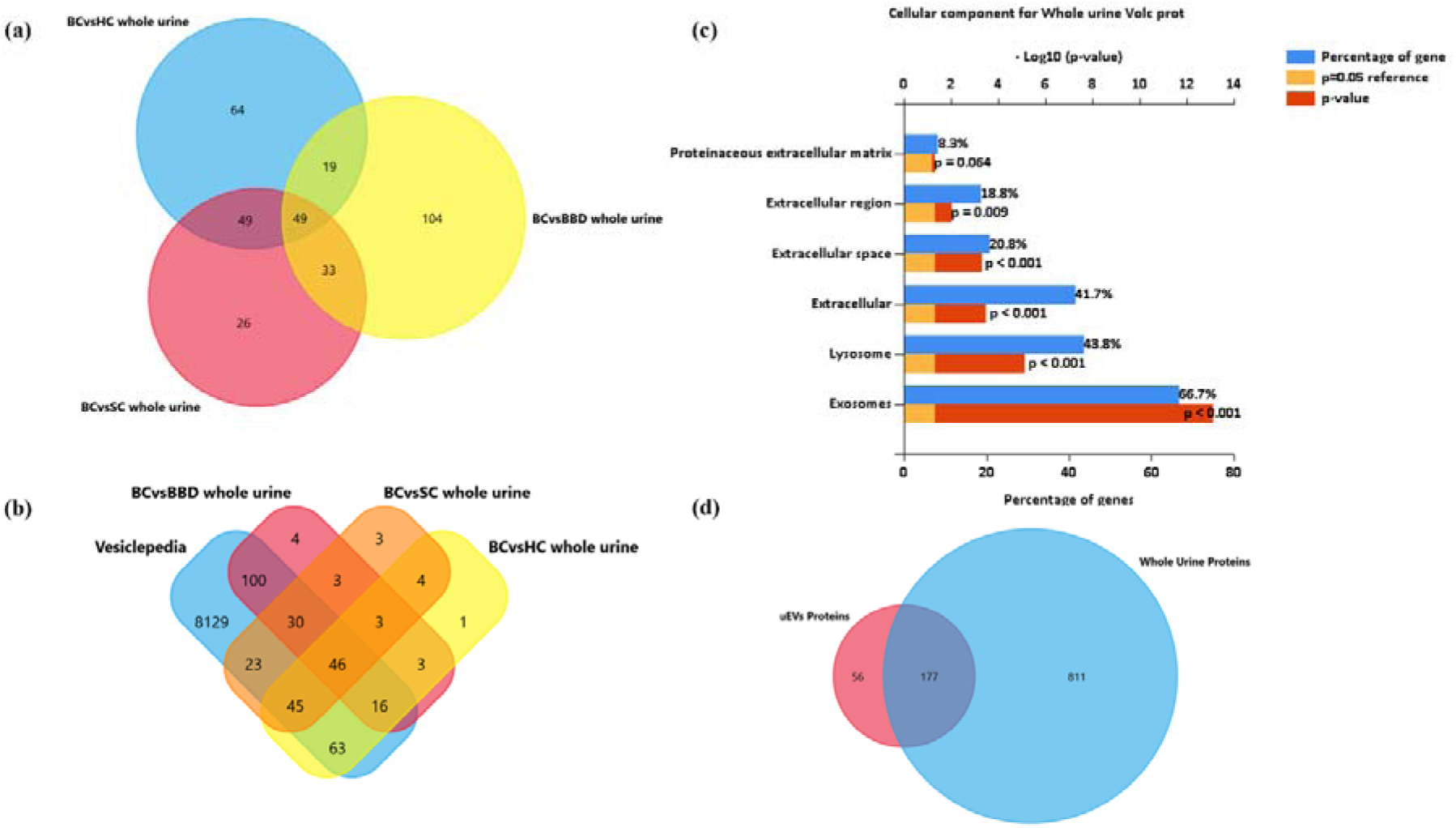
D**i**fferentially expressed proteins (DEP) in the urine of breast cancer (BC). Venn diagrams of **(a)** DEF of BC versus benign breast disease (BBD), symptom controls (SC), and healthy controls (HC) and **(b)** Overlap of DEF with entries on the Vesiclepedia Database. **(c)** Predicted Cellular locations of BC DEPs. Note the predominance of exosomes as the location of the DEPs. **(d)** Venn diagram of proteins identified in the whole urine and the uEV proteomes.

The likelihood of these DEPs being associated with EVs, was initially assessed by comparison with the Vesiclepedia database (Figure 1b). Among the DEPs common to all comparisons, 46 were predicted to be EV-associated. Additionally, some DEPs specific to individual comparisons were also predicted to be EV-associated, including 100 DEPs from the BC vs BBD comparison. Considering the predicted cellular locations of the DEPs, most were associated with exosomes and lysosomes (Figure. 1c). A total of 66.7% of these genes were significantly (< 0.001), linked to exosomes and 43.8% to lysosomes, This confirmed the findings of our earlier study (Zainurin et al., 2025) and served as the foundation of our subsequent uEV proteome focused assessment.

### Proteomic Profiling of urinary Extracellular Vesicles (uEVs)

To directly investigate the uEV proteome, uEVs were isolated from the same urine samples used to derive the whole urinary proteome. To assess the successful enrichment of uEVs, transmission electron microscopy (TEM) was employed to examine particle size and morphology (Figure 2). TEM analysis revealed predominantly spherical, membrane-bound vesicle-like structures with diameters ranging approximately from 100 nm to 500 nm, consistent with the expected size range and morphology of EVs. The observed vesicle-like particles were morphologically comparable to EVs reported previously using TEM in other biofluids, including serum EVs described by Buschmann et al. (2018), supporting their classification as extracellular vesicle–like structures (Buschmann et al., 2018). The presence of protein cargo associated with the isolated uEVs was confirmed by SDS-PAGE (data not shown).

**Figure 2:**
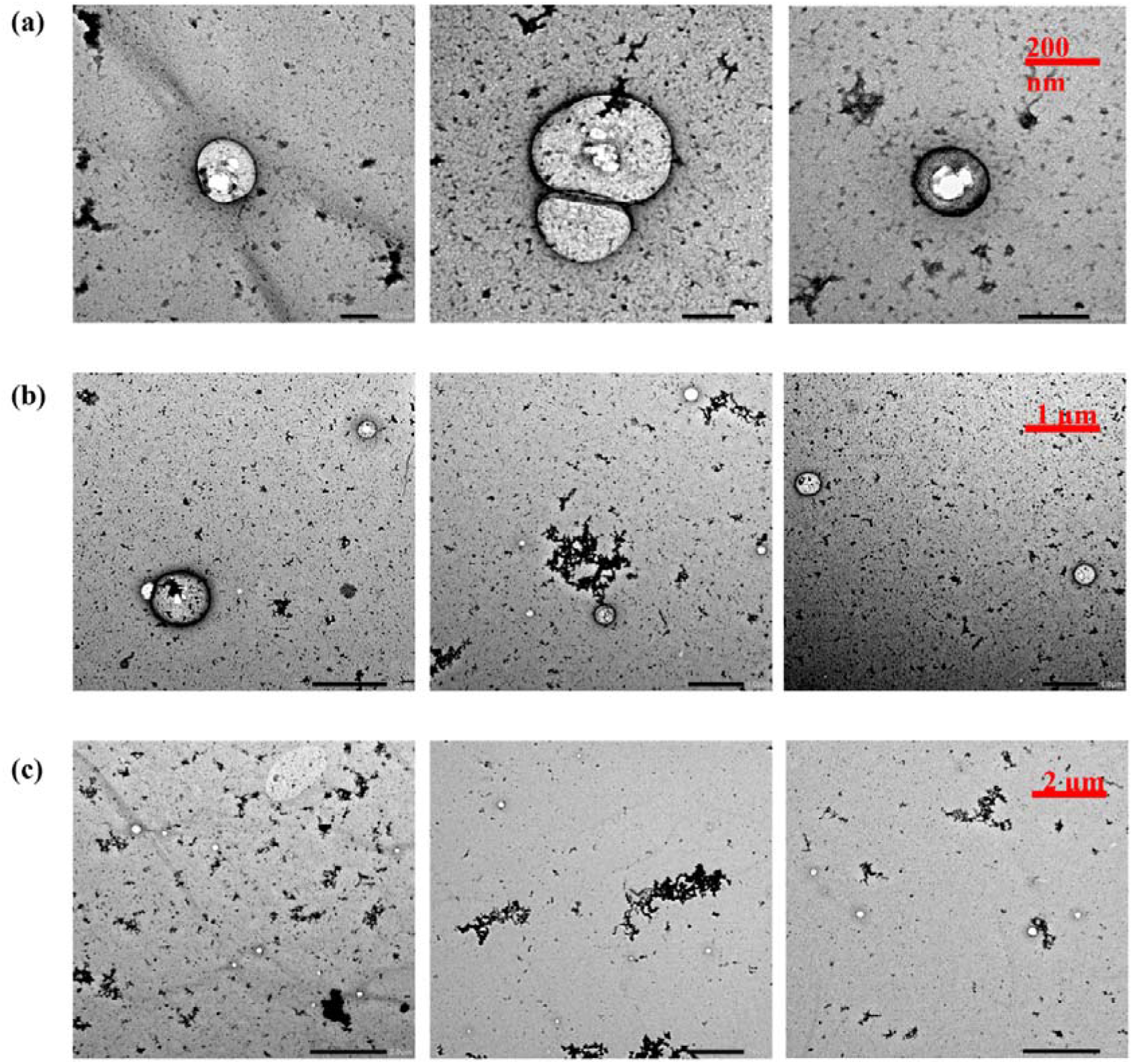
Imaging urinary extracellular vesicles (EV) by transmission electron microscopy. Differently sized EVs **are** shown based on imaging at **(a)** 200 nm **(b)** 1 µm and **(c)** 2 µm scales.

The uEV proteome was screened and a total of 257 unique proteins were identified across all the sample sets (Table S2). To unambiguously link these proteins to EVs, these were compared to the ExoCarta database Top 100 proteins often identified in EVs of whatever origin (http://exocarta.org/sEV_top100). Fifteen proteins of our uEV set were listed amongst the top 100 EV proteins list. These included *Programmed Cell Death 6 Interacting Protein* (PDCD6IP), *Glyceraldehyde-3-Phosphate Dehydrogenase* (GAPDH), *Syntenin-1* (SDCBP), *Annexin A2* (ANXA2) and *Actin, cytoplasmic 1* (ACTB). PDCD6IP and SDCBP in particular are considered as canonical EV markers as recommended by the MISEV guidelines (Minimal Information for Studies of Extracellular Vesicles) (https://isevjournals.onlinelibrary.wiley.com/doi/10.1002/jev2.12404). Further assessments suggested that 177 uEV proteins (76%) had been previously detected in the whole urine proteome. Thus, 56 uEV proteins (24 %) appeared only in the uEV proteome (Figure. 1d)

### Multivariate Analysis of the uEV Proteome

PCA did not reveal distinct uEV proteomic clustering between the four sample classes (BC, BBD, SC and HC), although some separation was observed with BBD, suggesting that major sources of variance were not linked to class differences (Figure 3a). In contrast, supervised PLS-DA showed a clear separation between the classes (Figure 3b). Cross-validation, of the model suggested an accuracy of 0.80, R² = 0.996, and Q² = 0.65, demonstrating strong explanatory and predictive performance without evidence of overfitting.

**Figure 3:**
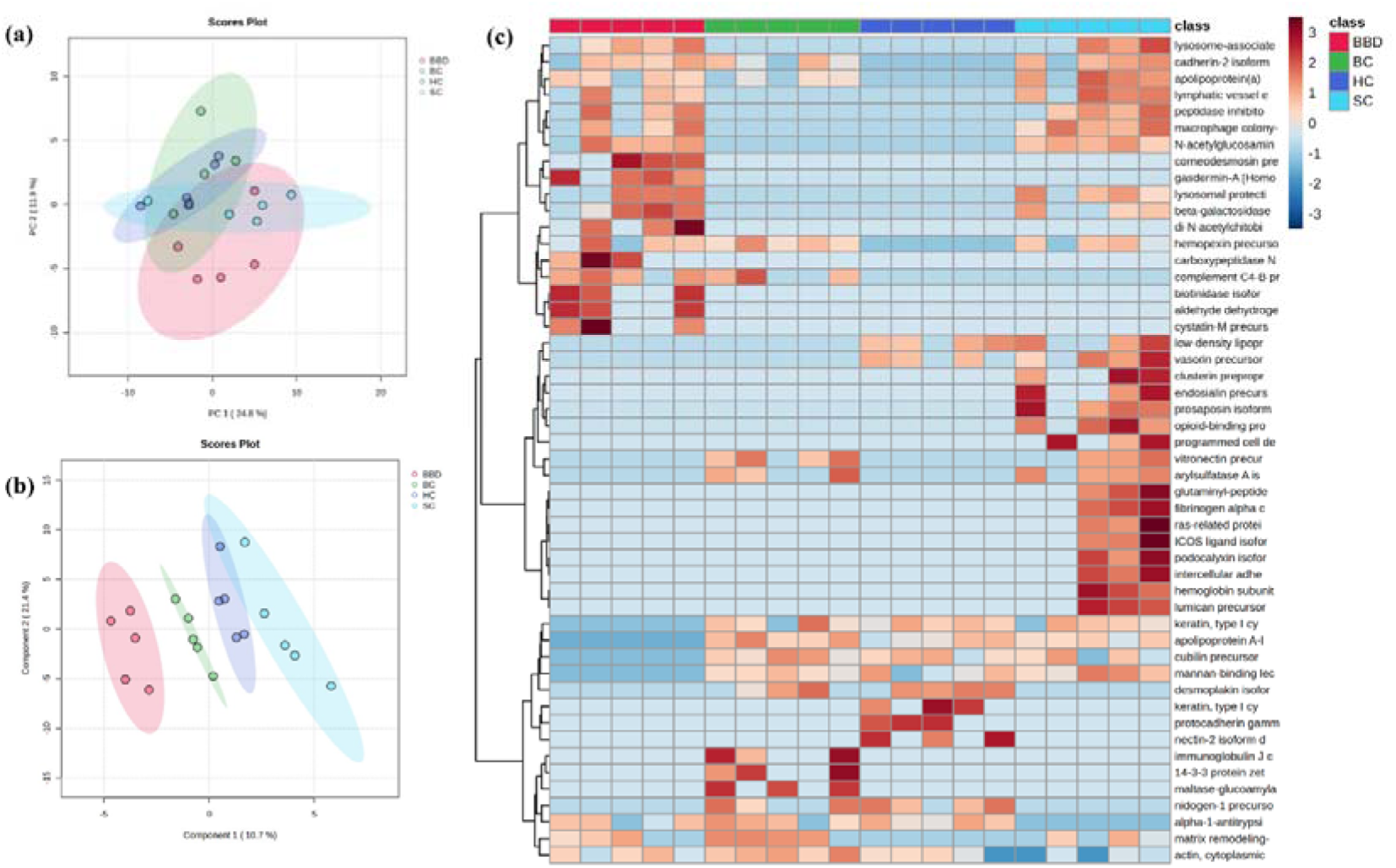
Identifying breast cancer (BC) associated changes in the urinary extracellular vesicle (EV) proteome. **(a)** Principal component analysis (PCA) and **(b)** partial least squared-discriminant analysis of urinary EV of BC, benign breast disease (BBD), symptom controls (SC), and healthy control (HC) groups. **(c)** Heatmap of differentially expressed proteins (DEP) across all groups.

The major sources of variation were identified by ANOVA (*P* <0.05, correcting for false discovery rates [FDR]) and displayed on a heat map (Figure 3b). This suggested that the BBD and SC groups were the most distinctive groups when compared with HC at the uEV level. However, there were three proteins that were highly expressed in breast cancer group but absent in the other groups: namely immunoglobulin J chain (JCHAIN), maltase-glucoamylase 2 (MGAM2) and 14-3-3 protein zeta/delta (YWHAZ).

### Pairwise comparisons of uEV Proteomes Between Breast Cancer, Benign Breast Disease, Symptom Controls and Healthy Controls

Pairwise comparison of the BC vs BBD classes using volcano plots (*P* <0.1/ fold change ≥ 2) targeted 40 DEPs (Figure 4a). Log_2_ fold change values suggested that 16 uEV proteins were upregulated and 24 downregulated in the BC group. DEP up and down regulated in BC were assessed using Metascape to link these to discrete biological pathways (Figure 4b). Significantly enriched terms in the BC upregulated DEP list were “positive regulation of cell-substrate adhesion” (GO:0010811), “homotypic cell-cell adhesion (GO:0034109) and humoral immune response” (GO:0006959). BC downregulated DEPs were enriched in “aminoglycan catabolic process” and “diseases of metabolism”.

**Figure 4:**
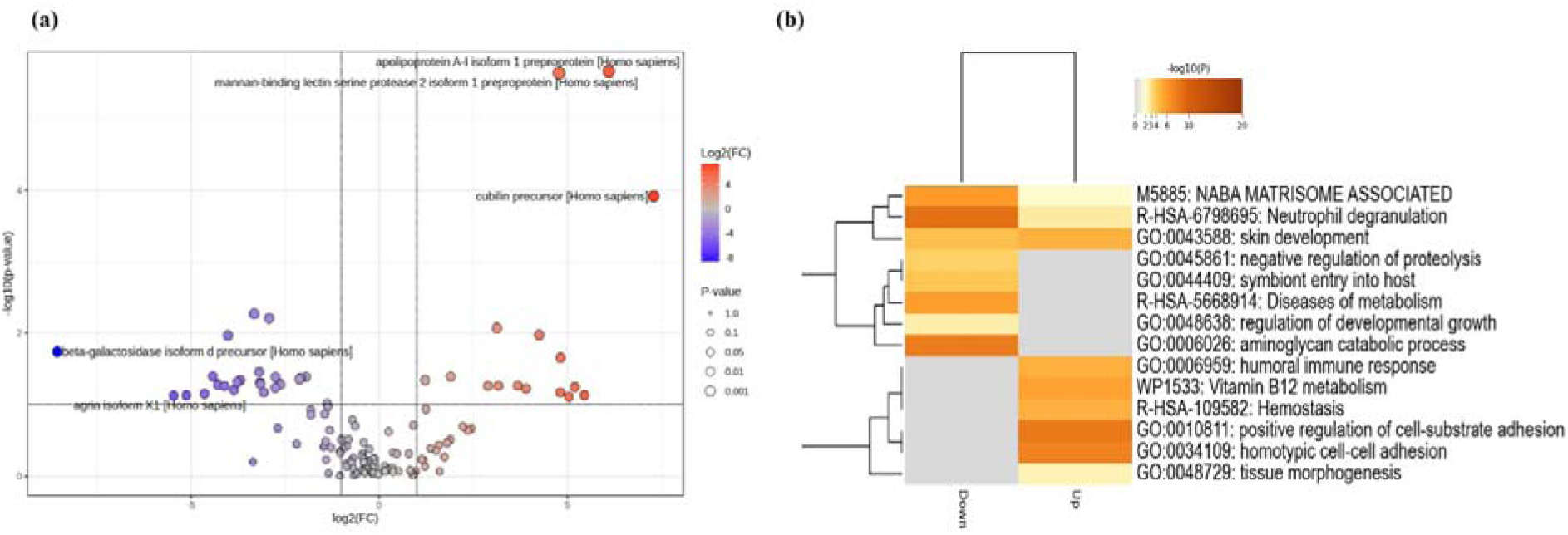
Differentially expressed proteins (DEP) in the urinary extracellular vesicle (EV) proteomes between breast cancer (BC) and benign breast disease (BBD) samples. DEP identified by **(a)** volcano plots. **(b)** A heatmap of DEP showing the functional terms associated with each DEP.

The volcano plots for pairwise comparison between BC and SC revealed 45 DEPs where 14 were upregulated and 31 downregulated in the BC group (Figure 5a). The enrichment analysis suggested that the upregulated proteins showed strong enrichment in “peptide cross-linking” (GO:0018149) and “cytoskeletal organization” (hsa04820), “post-translational protein phosphorylation” (R-HSA-8957275), and “vesicle-mediated transport” (R-HSA-5653656) Down regulated proteins targeted “Integrin cell surface interactions” (R-HSA-216083), “regulation of hydrolase activity” (GO:0051336) and “lysosomes” (hsa04142) (Figure 5b).

**Figure 5:**
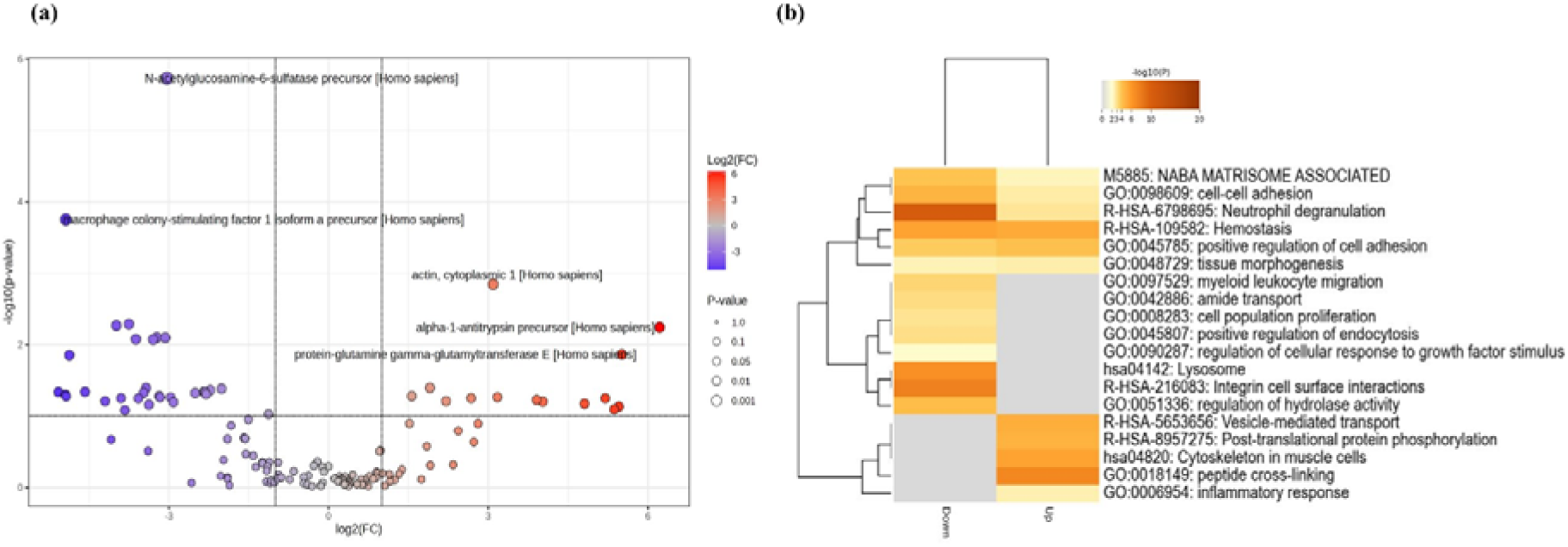
Differentially expressed proteins (DEP) in the urinary extracellular vesicle (EV) proteomes between breast cancer (BC) and symptom control (SC) samples. DEP identified by (a) volcano plots. (b) A heatmap of DEP showing the functional terms associated with each DEP.

In a comparison between BC and HC, 11 DEPs were up-regulated and 26 down-regulated. Enrichment analysis for these proteins suggested the up-regulated DEP were involved in “scavenging of heme from plasma” (R-HAS-2168880), “hemostasis” (R-HAS-109582), “complement system” (WP2806) and “post translational protein phosphorylation” (R-HAS-8957257). Downregulated DEPs were associated with “neutrophil degranulation”, “cell-cell adhesion” and “morphogenesis of an epithelium” (Figure 6).

**Figure 6:**
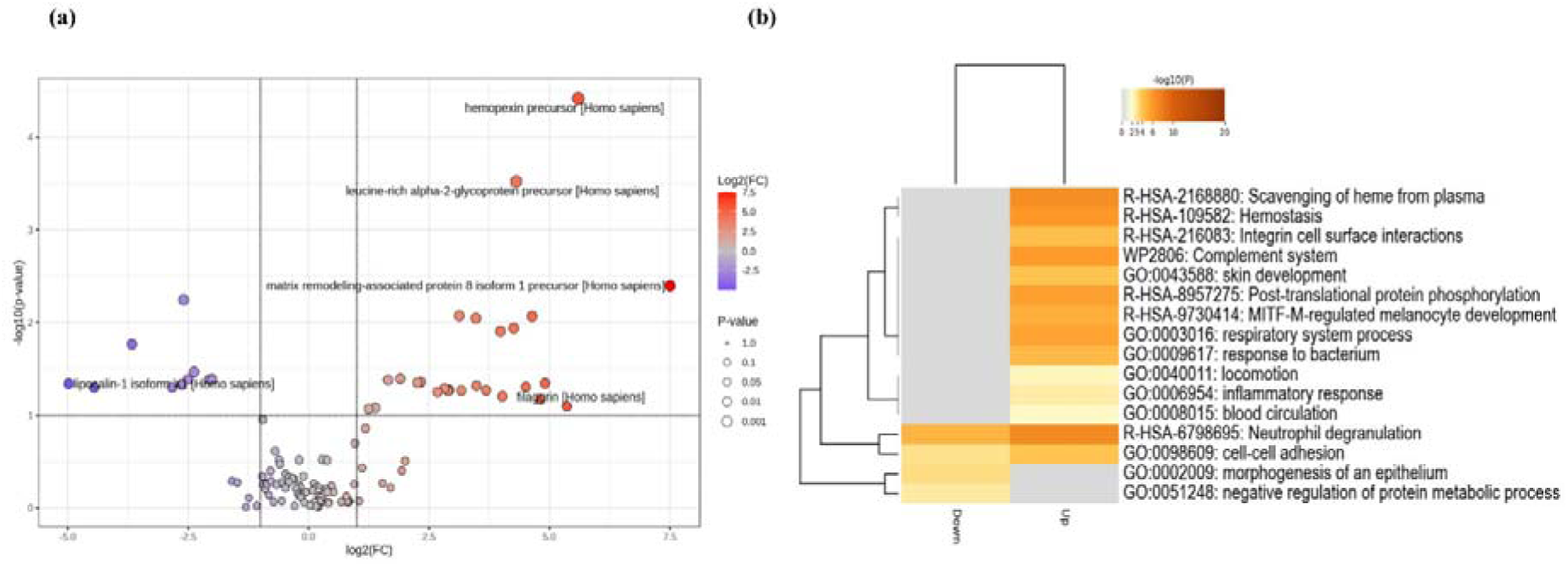
Differentially expressed proteins (DEP) in the urinary extracellular vesicle (EV) proteomes between breast cancer (BC) and healthy control (HC) samples. DEP identified by **(a)** volcano plots. **(b)** A heatmap of DEP showing the functional terms associated with each DEP.

Overlap between the uEV DEPs targeted by pairwise comparisons was show in a Venn diagram (Figure. 7a). This indicated seven proteins were consistent across all comparisons, namely, SERPINB1, LCN1, PTP1B, ACTB, YWHAZ, JCHAIN and APOA1.

**Figure 7:**
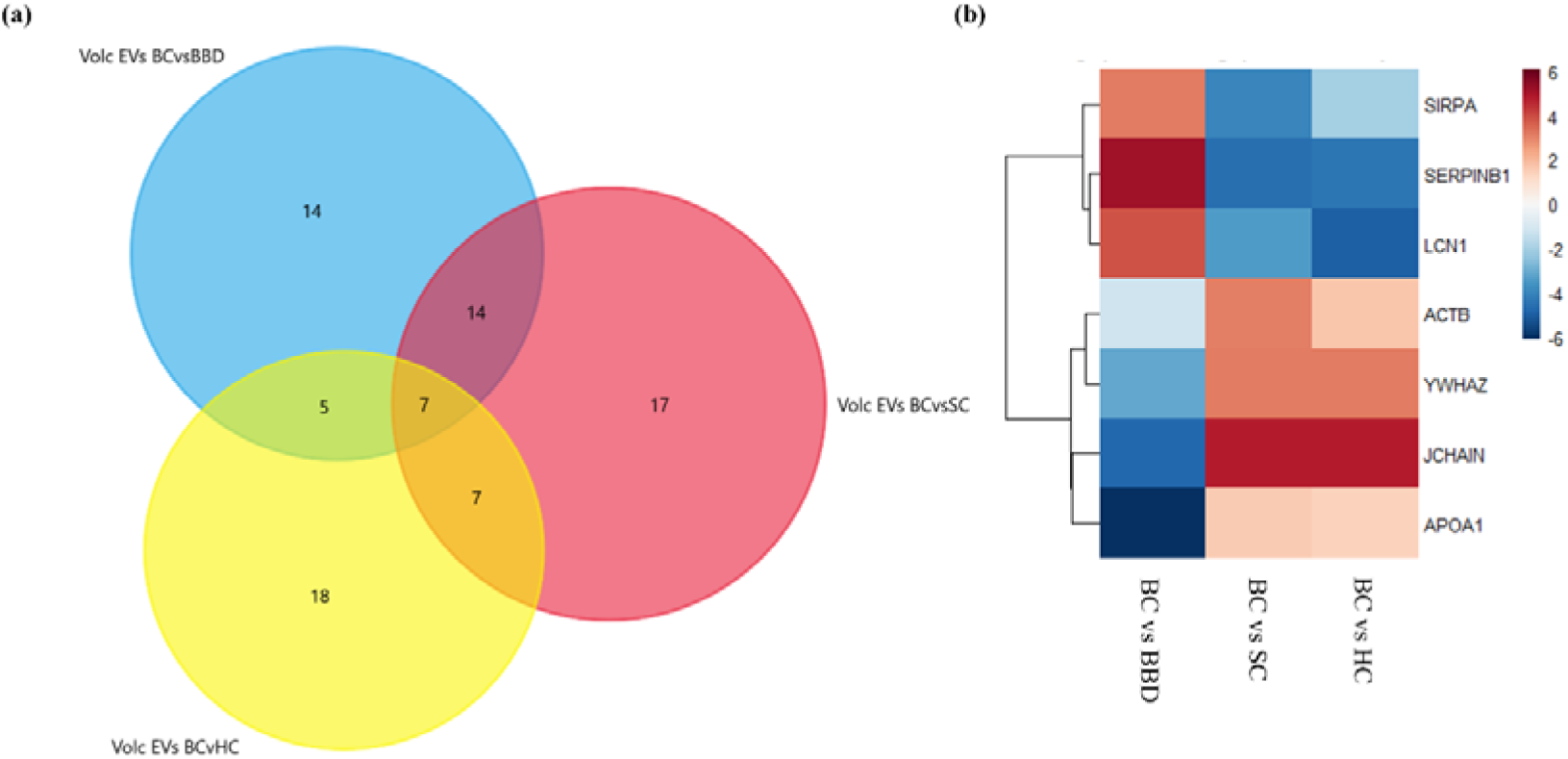
DEPs identified by pairwise comparisons between breast cancer (BC) versus benign breast disease (BBD), symptom controls (SC), and healthy controls (HC) as shown by **(a)** a Venn diagram. The seven common DEP in each comparison are) SERPINB1 — Serpin family B member 1, LCN1 — Lipocalin 1, SIRPA — Signal regulatory protein alpha, ACTB — Actin, beta, YWHAZ —Tryptophan 5-monooxygenase activation protein zeta, Ig JCHAIN and APOA1 — Apolipoprotein A1. The relative expression of each of the seven common DEPs in each pairwise comparison is shown by a **(b)** heatmap based on Log_2_ (Fold Changes)

To further assess these proteins potential in separating between breast cancer and the other groups, we preformed univariate ROC curve analysis for each comparison (Table 1). All seven proteins exhibited AUC values greater than 0.8. Considering whether the DEPs were either up or down regulated compared to BC, it was noted that there were consistent changes with SC and HC, but these were always the opposite of the BC vs BBD comparison (Figure. 7b).

**Table 1:**
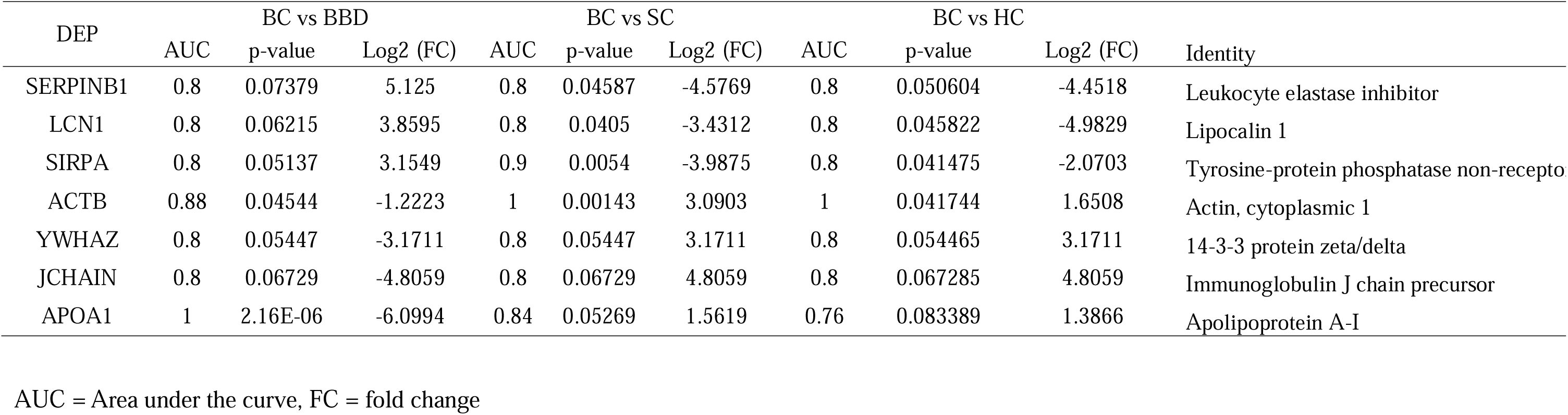
Key differentially expressed proteins (DEP) in Breast Cancer (BC) compared to Benign Breast Disease (BBD), Symptom controls (SC) and Healthy Controls (HC) in the urinary extracellular vesicle proteome (uEV).

## Discussion

This study builds upon previous work aimed at identifying biomarkers for breast cancer in whole urine samples (Zainurin et al., 2025). This previous examination described five potential urinary protein biomarkers that could distinguish between BC and BBD, SC and HC with an AUC of 0.985, 0.989 and 0.999, respectively. In the current study, we applied a similar analytical workflow and observed that several proteins discriminating between BC, BBD, SC and HC, to explore how urinary DEPs could be associated with uEVs.

We first sought to substantiate our hypothesis that DEPs associated in BC in the whole urinary proteome were uEV associated. This involved partially repeating our earlier study (Zainurin et al., 2025) and bioinformatically relating these to EV protein databases. This successfully suggested that the DEPs included EV located proteins. To experimentally establish an EV localisation of some DEPs, uEVs were isolated from the same urine samples. EV’s were effectively isolated as indicated by TEM and EV proteome database comparisons of the newly isolated proteins. Of note, the targeted uEV DEPs were also relevant to BC tumours.

BC vs. BBD DEPs included proteins involved in modifying cell interactions (positive regulation of cell-substrate adhesion, homotypic cell-cell adhesion, aminoglycan catabolic processes) and immune responses (humoral immune response); both features that are important in cancer. Cell–substrate adhesion enhances the ability of BC cells to interact with the ECM, primarily through integrins and focal adhesion complexes. Integrins mediate adhesion between cells and the ECM, the lower levels suggest reduced adhesion signalling, which could influence tissue integrity and cell communication (Hamidi & Ivaska, 2018). Further, integrins regulate breast cancer cell adhesion, migration, ECM remodelling, angiogenesis, and resistance to therapy (Yousefi et al., 2021). Integrin interactions activate downstream signalling pathways such as FAK, Src, PI3K, and Rho/ROCK, which collectively promote cell survival, proliferation, mechanotransduction, and migration. Aminoglycan changes could also breakdown glycosaminoglycans (GAGs), such as hyaluronan, heparan sulfate, and chondroitin sulfate, which are major components of the ECM (Heldin et al., 2013). Homotypic cell–cell adhesion mediated mainly by E-cadherin and associated catenins and their disruption leads to EMT, a key feature in BC metastasis (Chong et al., 2024). In BC the activity of tumour-infiltrating B cells (TIL-B) and the antibodies they produce are important components of the humoral immune response. Increases in TIL-B can lead to increased BC invasive potential (Garaud et al., 2019).

Considering BC DEP with the control groups (SC and HC), these also suggest changes in the ECM. This mostly clearly indicated by pathways that the BC vs. SC DEPs were associated with linked to integrin cell surface interactions, regulation of hydrolase activity and lysosomes. Altered lysosomal and hydrolase-related proteins may indicate shifts in degradative and metabolic processes, which are often associated with cancer progression (Kallunki et al., 2013; Perera & Zoncu, 2016). Upregulated DEPs showed enrichment in “peptide cross-linking” and “cytoskeletal organization” both suggestive of structural changes (Aseervatham, 2020), whilst post-translational protein phosphorylation and vesicle-mediated transport indicate altered signalling and trafficking processes inside cells (Gorshtein et al., 2021). The BC vs HC DEPs were linked to “neutrophil degranulation”, “cell-cell adhesion” and “morphogenesis of an epithelium”, suggesting reduced innate immune activity and impaired epithelial organization and adhesion, which are often linked to tissue remodelling and cancer progression (Bruner & Derksen, 2018; Santarosa & Maestro, 2021).

Considering how these enhanced pathways reflected changes in individual proteins, seven were shown to be consistently altered in every comparison with BC. Of interest, MGAM2 and YWHAZ have been shown to have higher expression in cancer tissues compared to normal tissues (Mei et al., 2020; Xu et al., 2019). YWHAZ, which encodes the 14-3-3ζ protein, is widely recognized as an oncogenic driver in multiple cancers, including BC where it is an indicator of a poor prognosis. YWHAZ interacts with DAAM1, a cytoskeletal regulator, to promote cell migration and metastasis (Mei et al., 2021). YWHAZ is described as a multifunctional signalling protein involved in pathways such as PI3K/AKT, epithelial–mesenchymal transition, and chemotherapy resistance (Gan et al., 2020). In contrast, no mechanistic or clinical studies have shown a role for MGAM2, a putative maltase-glucoamylase, in breast tumor biology. JCHAIN is a linker protein of multimeric antibodies IgA and IgM and lower levels could reflect immunosuppression, possibly in the tumour microenvironment serving as a potential prognostic marker (Shi et al., 2025; Zhao et al., 2025). However, we observed increases in the levels of JCHAIN in uEVs in the BC group.

SERPINB1 is an intracellular protein primarily expressed in neutrophils and known for its role in inhibiting proteases to regulate immune responses and preventing tissue damage (Baumann et al., 2013). In breast cancer SERPINB1 has been linked with cancer metastasis and proposed as a potential biomarker of poor prognosis (Mani, 2015). LCN1 is increasingly recognized as a significant biomarker for breast cancer with high expression associated with poor prognosis and reduced disease-free survival in all types of breast cancer (Yang et al., 2019; Zhang et al., 2020).

SIRPA is transmembrane protein primarily expressed on macrophages and dendritic cells, playing a crucial role in immune response. SIRPA interacts with CD47 expressed on normal cells to prevent phagocytosis, hence modulating the immune system. In cancer, malignant cells tend to express CD47 to evade detection and destruction by the immune system (Takahashi, 2018). While evidence suggest that CD47/SIRPA interaction is a sign for cancer progression and immune evasion, the use of SIRPA expression alone remains limited and warrants further investigation. APOA1 is a major protein component of high-density lipoproteins (HDL), playing a key role in lipid transport and anti-inflammatory processes. Contrary to our observation of elevated APOA1 in uEVs from the BC group, previous studies have reported reduced circulating APOA1 levels in breast cancer patients, with lower plasma APOA1 being associated with an increased risk of breast cancer development (Darwish et al., 2023). This discrepancy may be explained by differences in sample type, as previous studies measured circulating APOA1 in plasma, whereas the present study assessed EV-associated APOA1 in urine. EVs are known to selectively package proteins, and it is possible that APOA1 is enriched within vesicles in breast cancer despite reduced circulating levels. This may reflect a redistribution of APOA1 into EVs or distinct functional roles of EV-associated APOA1 in tumour-related processes.

ACTB is a fundamental cytoskeletal protein essential for maintaining cell structure, motility, and intracellular transport and is involved in EMT. Pan-cancer analyses indicate that ACTB is frequently upregulated across multiple tumour types, suggesting its role in promoting cytoskeletal dynamics and potentially serving as a marker of aggressive disease (Gu et al., 2021).

Our observations indicate that the uEV populations can reflect changes that are commonly seen in tumour cells or in the TME, for example, ECM changes. In addition, they may also reflect potential roles for EVs in facilitating metastasis. For example, SERPINB functions as an inhibitor of granzyme B produced by natural killer (NK) cells and cytotoxic T lymphocytes allow the evasion of the immune responses and contributes to cancer progression (Wang et al., 2021). Such observations suggest a continuity between blood/lymphatic EVs and the uEV population (Iannotta et al., 2024). Most uEVs arise from the kidney and urinary tract epithelium, particularly from the apical membranes of nephron and collecting-duct cells as urine flows through the nephron (Grange et al., 2023). However, our observations would suggest that some EVs arise from the tumor environment itself. We did not assess the presence of our uEV proteome changes in blood EV but clearly that is an important future objective.

These findings reinforce the hypothesis that uEV proteomics represents a promising avenue for biomarker discovery, given the role of EVs in intercellular communication and their enrichment in disease-specific cargo. The seven identified proteins are promising candidates for breast cancer biomarkers; however, additional validation is essential. Future studies should include larger numbers of participants and incorporate blinded testing, where the disease status of participants is unknown during analysis, to evaluate whether these proteins can accurately predict breast cancer prior to confirmation by clinical diagnosis.

## Conflict of Interest

The authors declare no conflict of interest.

## Funding Information

N.L. is partially supported by Institute of Biomedical Science grant (IBMS)(Urinary protein extracellular vesicles as a source of biomarkers for early detection of breast cancer) grant. N.A.A.Z is supported by a Graduate Excellence Programme (GrEP), Majlis Amanah Rakyat (MARA) fellowship (Grant No. 330408376064). L.A.J.M is partially supported by Shandong Province “Double-Hundred Talent Plan” Teams (Grant No. WSR2023049). R.M.M. acknowledges grant funding (Welsh Government: Establishing Cutting Edge Veterinary Research Laboratories for Wales; European Development Regional Fund Sêr Cymru program Grant - 80761-AU185) for the acquisition of the Ultimate 3000 RSL Nano with ES082 EASY-Spray Source for mass spectrometry.

## Ethics Statement

The study was reviewed and approved by Health Research Authority (HRA) and Health and Care Research Wales (HCRW). All participants involved provided informed consent prior to participation in the study.

## Registry and the Registration No. of the study

IRAS Project ID: 306872; Protocol no: AU/IBERS/010; REC reference: 21/SC/0411; CPMS study ID: 54143.

## Data Availability Statement

Proteomic data has been deposited in the PRIDE database under accession number PXD072769.

## Notes

### Competing Interest Statement

The authors have declared no competing interest.

### Author Declarations

Ethical approval for the BECA (OMICs Approaches to Improve the Diagnosis, Management and Treatment of Breast Cancer) clinical project was obtained from the Health Research Authority (HRA) and Health and Care Research Wales (HCRW) IRAS Project ID: 306872 and CPMS Study ID: 54143.

